# Computational systematics of nutritional support of vaccination against viral and bacterial pathogens as prolegomena to vaccinations against COVID-19

**DOI:** 10.1101/2021.09.10.21263398

**Authors:** Ivan Y. Torshin, Olga A. Gromova, Alexander G. Chuchalin

## Abstract

A total of 6,628 PUBMED-registered publications on the relationships between the effects of vaccination and the provision of micronutrients have been studied by methods of topological analysis of text data. In case of insufficient intake of certain micronutrients, the functioning of the acquired immunity is disrupted resulting in an imbalance of populations of T-cells CD4+/CD8+ and of B-lymphocytes. Nutritional supplements of folate, vitamins A, D and B12, which are recognized regulators of cell division, support a wide range of lymphocyte populations. Trace elements zinc, iron, selenium, manganese and omega-3 polyunsaturated fatty acids are also important for supporting the mechanisms of acquired immunity. The data presented show that a course intake of these micronutrients by patients planning vaccination can significantly improve its effectiveness. In particular, these micronutrients can increase the titers of antibodies to pathogens, and to reduce the percentage of patients who still contract infection after vaccination. Supplements of these micronutrients can also contribute to the safety of vaccination: to prevent malaise and, in the unfortunate case of contracting infection despite the vaccine, to reduce the severity of the course and the mortality from the corresponding infection.

## Introduction

Vaccination against specific strains of viral and bacterial pathogens is considered to be the most effective approach to anti-infective protection at population level. Vaccination aims to activate acquired immunity, which implies the production of pathogen-specific antibodies in sufficient concentration. At the same time, vaccination does not stimulate and does not support the innate antiviral and antibacterial immunity [1, 2].

Fundamental issues of interactions between acquired and innate immunity become particularly relevant in viral epidemics and especially in pandemics [3]. For example, an analysis of molecular biological pathways involved in innate immunity against the SARS-CoV-2 [4] showed that an increase in the body’s supply of vitamins A, D, magnesium, zinc can be an important and yet inexpensive resource for enhancing the activity of the innate immunity (interferon system, in particular) against RNA viruses [4]. These micronutrients and a number of pharmaceuticals can show a direct antiviral effect, inhibiting viral replication in cell cultures [5].

The augmentation of the supply of micronutrients to the general population is also important to prevent chronic comorbid pathologies, which typically involve numerous micronutrient insufficiencies [6]. Population-wide vitamin and trace element deficiencies and insufficiencies represent a pressing problem in various branches of medicine (more details at www.trace-elements.ru). Large-scale clinical and epidemiological studies conducted in Russia and abroad indicate quite high occurrences of micronutrient deficiencies and their relationship with numerous chronic pathologies, including impaired skin barrier function, impaired immunity [7] and chronic inflammation [8]. In children of various age groups the supply of micronutrients is not better either (50% on average per micronutrient) and vitamin insufficiencies were associated with decreased activity of detoxification systems, increased frequency of asthma attacks and decreased immunity to common cold [9].

The data of preclinical and clinical studies suggest that micronutrient deficiencies adversely affect the functioning of the acquired immunity and, therefore, negatively affect the effectiveness and the safety of vaccination, including vaccines against various RNA viruses (influenza viruses, measles, RS virus, coronaviruses etc.). For example, supplementation of vitamins A and D during immunization with influenza virus vaccine improved the weak antibody response in the mucous membrane of vitamin-deficient mice [10]. Vitamin supplementation to mice during vaccination against pneumococcus increased immunogenicity and survival after infection with Streptococcus pneumoniae [11]. Note that a number of vitamins and microelements are used as adjuvants for vaccines that enhance the acquired immune response: selenium nanoparticles [12], iron nanoparticles [13] and calcium carbonate [14], vitamins A, E and catechins [15].

The issues of improving the efficiency and the safety of vaccination are very relevant in the case of coronavirus infection COVID-19. Currently, there is no reliable scientific data on the full range of consequences from the use of certain vaccines against COVID-19 among the general population. However, separate news reports suggest that the efficacy and safety of COVID-19 vaccination may decline sharply in the presence of concomitant micronutrient deficiencies. For example, the elderly are prone to form multiple micronutrient deficiencies. A news item succinctly reported that after the COVID-19 vaccination campaign in a Spanish nursing home, all 78 residents fell ill. Seven of them died, and four were hospitalized [16]. As a result of the use of the same vaccine in Norway, 23 people died [17] (mainly elderly patients). So, early compensation of micronutrient deficiencies can be a promising direction for increasing the effectiveness and safety of vaccination against COVID-19.

This work presents the results of a systematic computer analysis of studies on the relationships between the supply of vitamins and microelements and the functioning of the acquired immunity (thus the effectiveness and safety of vaccination against viral and bacterial pathogens).

## Materials and methods

The publications on the relationship between the effects of vaccination and the provision of various micronutrients were found by using the request “(vaccine OR vaccination) AND (folic OR folate OR riboflavin OR niacin OR nicotinamide OR pantothenic OR pyridoxine OR myoinositol OR biotin OR cyanocobalamin OR vitamin OR polyunsaturated-OR PUFA 3 OR zinc OR selenium OR magnesium OR iodine OR copper OR manganese OR calcium OR iron OR lithium)” to PUBMED database (6,628 publications on February 2021). This array of publications was analyzed using topological [18] and metric approaches to data analysis [19, 20].

Since among publications on vitamins there is often an increased percentage of texts having a pronounced manipulative character [21], an array of 6,628 publications was checked by the ANTIFAKE system (www.antifake-news.ru). As a result, 521 publications with negative values of the beta-score were identified (see the description of the calculation procedure in [22]), i.e. publications in which manipulative content prevailed over the content. Typical examples of such publications have been texts describing attempts to use single-dose “megadoses” of vitamins (e.g., 50,000-100,000 IU of vitamin A, 100,000-500,000 IU of vitamin D, etc.). These studies were characterized by numerous violations of the fundamentals of pharmacology, biochemistry, data analysis and, due to an apparently low scientific quality, were excluded from further consideration.

## Results

In the course of a systematic analysis of the texts of 6,107 publications, 105 of the most informative biomedical terms were identified. These terms allow to distinguish the entire array of publications on the relationship of vaccination and micronutrients from a control sample of publications (6,100 articles randomly selected from 396,953 articles found by the query “(vaccine OR vaccination) NOT vitamin NOT polyunsaturated NOT PUFA NOT omega-3 NOT zinc NOT selenium NOT magnesium NOT iodine NOT copper NOT manganese NOT calcium NOT iron)”. Annotation of these terms in accordance with the international nomenclature of molecular biological processes (Gene Ontology, GO) made it possible to formulate a complex of molecular mechanisms describing the relationship between the effects of vaccination and micronutrients (Fig. 1).

**Figure 1.**
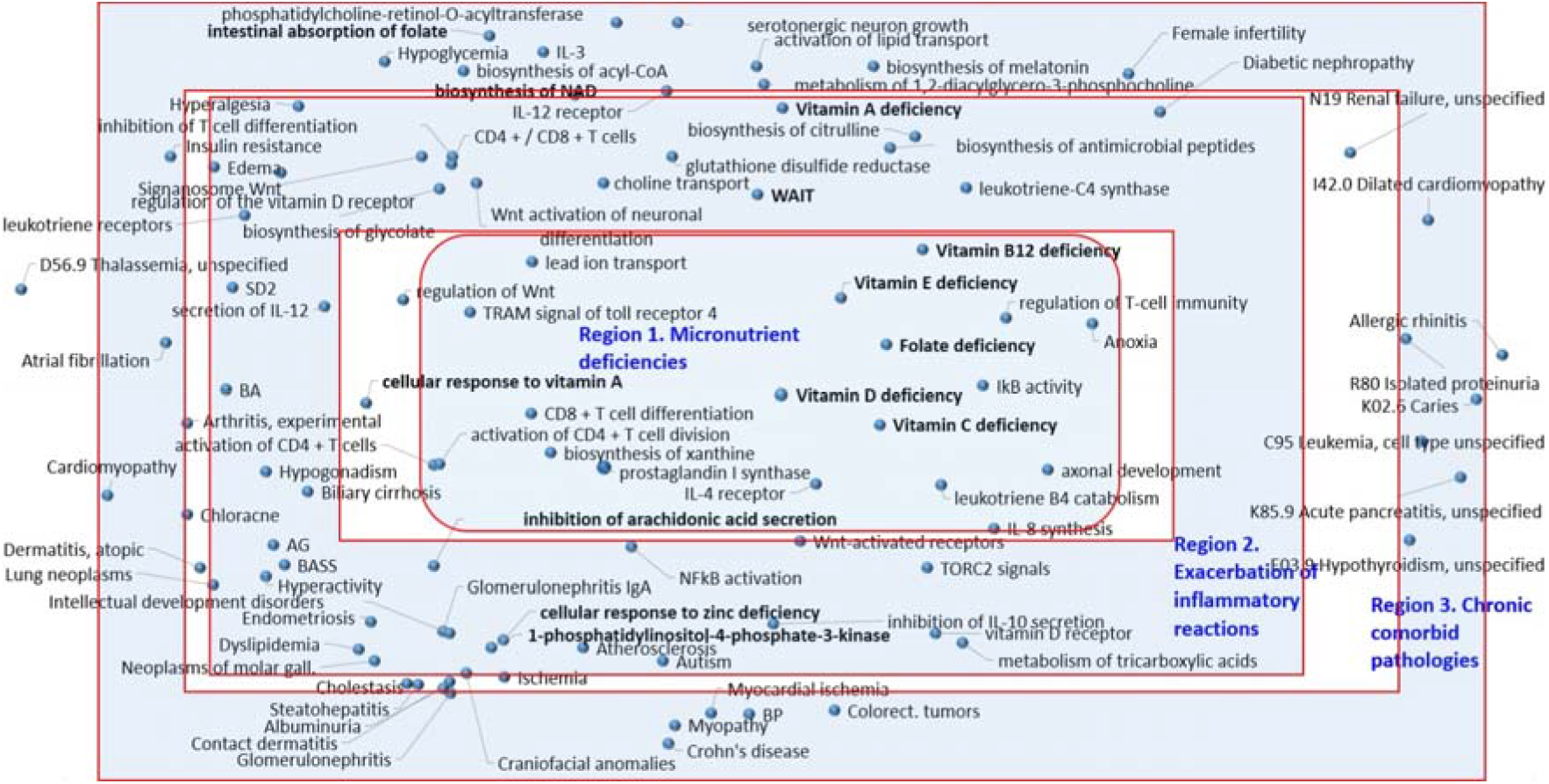
A metric diagram of the relationship between terms describing the effects of vaccination and terms indicating the availability of various micronutrients. The diagram was obtained as a result of a systematic computer analysis of 6,107 publications and represents an optimal projection of a multidimensional distance matrix configuration onto the plane. The distance between a pair of any points corresponding to a pair of terms is inversely proportional to the “interaction” of the terms (joint occurrence of terms in the studied sample of publications): the closer two arbitrary dots, the more often the joint use of terms occurs. Biological activities according to the international GO (Gene Ontology) nomenclature are shown in the figure without codes (GO codes are given in the text). Abbreviations: IDA, iron deficiency anemia; T2DM, diabetes mellitus; Hypertension, arterial hypertension; SLE, systemic lupus erythematosus; BA, Alzheimer’s disease; ALS, amyotrophic lateral sclerosis; PD, Parkinson’s disease.

Analysis of the diagram (Fig. 1) by the method of metric condensations [20] indicated the existence of the three regions of the diagram, to which the metric algorithms mapped the most informative biomedical terms. These terms represent a minimal set of the terms which allow a “deep learning” classification algorithm to distinguish between publications of the two classes with maximum accuracy [19] (namely, publications on relationships between vaccination and micronutrients and the publications the control set). It can be seen from Figure 1 that the three regions are concentric with the Region 1 “Micronutrient Deficiencies” being in the center of the diagram. The central location of the terms referring to micronutrient deficiencies indicates a close relationship with all other terms presented in the diagram [20]. In particular, the deficiencies of the micronutrients indicated on the diagram (vitamins A/D, group B, zinc, iron) are associated with exacerbation of inflammatory reactions, decreased activity of acquired immunity systems (Region 2) and with corresponding chronic comorbid pathologies (Region 3).

Compared to control publications (vaccination studies that did not study the effects of vitamins and trace elements), the terms reflecting disturbances of the acquired immunity, including cellular and humoral immunity, were significantly more often associated with publications on the relationship between vaccination and micronutrients. In particular, micronutrient deficiencies impair the regulation of T-cell immunity (GO term: 0002709), including T-helper cells and cytotoxic T-lymphocytes.

It is known that antigen-presenting cells (B-lymphocytes, macrophages, dendritic cells) cause activation (maturation) of T-helper CD4+ lymphocytes and cytotoxic CD8+ T-lymphocytes [1]. Micronutrient deficiencies (in particular, vitamin A deficiency) inhibit activation (GO:2000516 activation of CD4+ T cells) and division of T lymphocytes (GO:2000563 activation of CD4+ T cell division), which leads to dysfunction of T helper CD4+ lymphocytes.

At the same time, the functioning of both Th1 and Th2 T-helper responses is impaired. Recall that the Th1 response of T-helpers is associated with the synthesis of gamma-interferon, which activates defense systems against intracellular pathogens in bacterial and viral pathogens [2]. The Th2 response is associated with the production of IL-4 (GO:0004913 IL-4 receptor), the activation of eosinophils, and the switch of B-lymphocytes from the synthesis of antibodies of one class to the synthesis of antibodies of another class. Thus, a violation of the response of T-helpers leads to dysfunction of humoral immunity (including a decrease in the synthesis of antibodies by B-lymphocytes).

Micronutrient deficiencies can also lead to dysfunction of cytotoxic CD8+ T lymphocytes (GO:0043369 CD4+/CD8+ T cells, GO:0043374 CD8+ T cell differentiation, GO:0046639 inhibition of T cell differentiation), the activity of which is necessary to eliminate somatic cells infected by viruses.

Pro-inflammatory reactions provoked by vaccination are aggravated by micronutrient deficiencies since micronutrients are necessary to support the synergistic interaction of acquired immunity with the mechanisms of innate immunity [23] (GO:0002807 biosynthesis of antimicrobial peptides, GO:0035669 TRAM signal of toll receptor 4, GO:0004704 activation of NFkB, GO:0008384 IkB activity), normalization of the metabolism of pro-inflammatory: prostagland 1900139 inhibition of arachidonic acid secretion, GO:0031774 leukotriene receptors, GO:0036101 catabolism of leukotriene B4, GO:0004464 leukotriene-C4 synthase, GO:0008116 prostaglandin-I-synthase) and normalization of interleukin secretion (GO:2001180 secretion of IL-10, GO:0038156 IL-3, GO:0042228 synthesis of IL-8, GO:2001184 secretion of IL-12 etc).

These disturbances in the functioning of the immune system associated with micronutrient deficiencies (in particular, an increase in pro-inflammatory reactions) make a negative contribution to the pathophysiology of numerous chronic pathologies. These pathologies include osteoarthritis, atopic and contact dermatitis, glomerulonephritis, steatohepatitis, atherosclerosis, Crohn’s disease, and endometriosis. Mapping the most informative terms (Fig. 1) to the ICD-10 diagnoses showed that vaccination in persons with low micronutrient supply can also be associated with a more severe course of renal failure, proteinuria, hypothyroidism, pancreatitis, hemochromatosis and other pathologies (Fig. 2).

**Figure 2.**
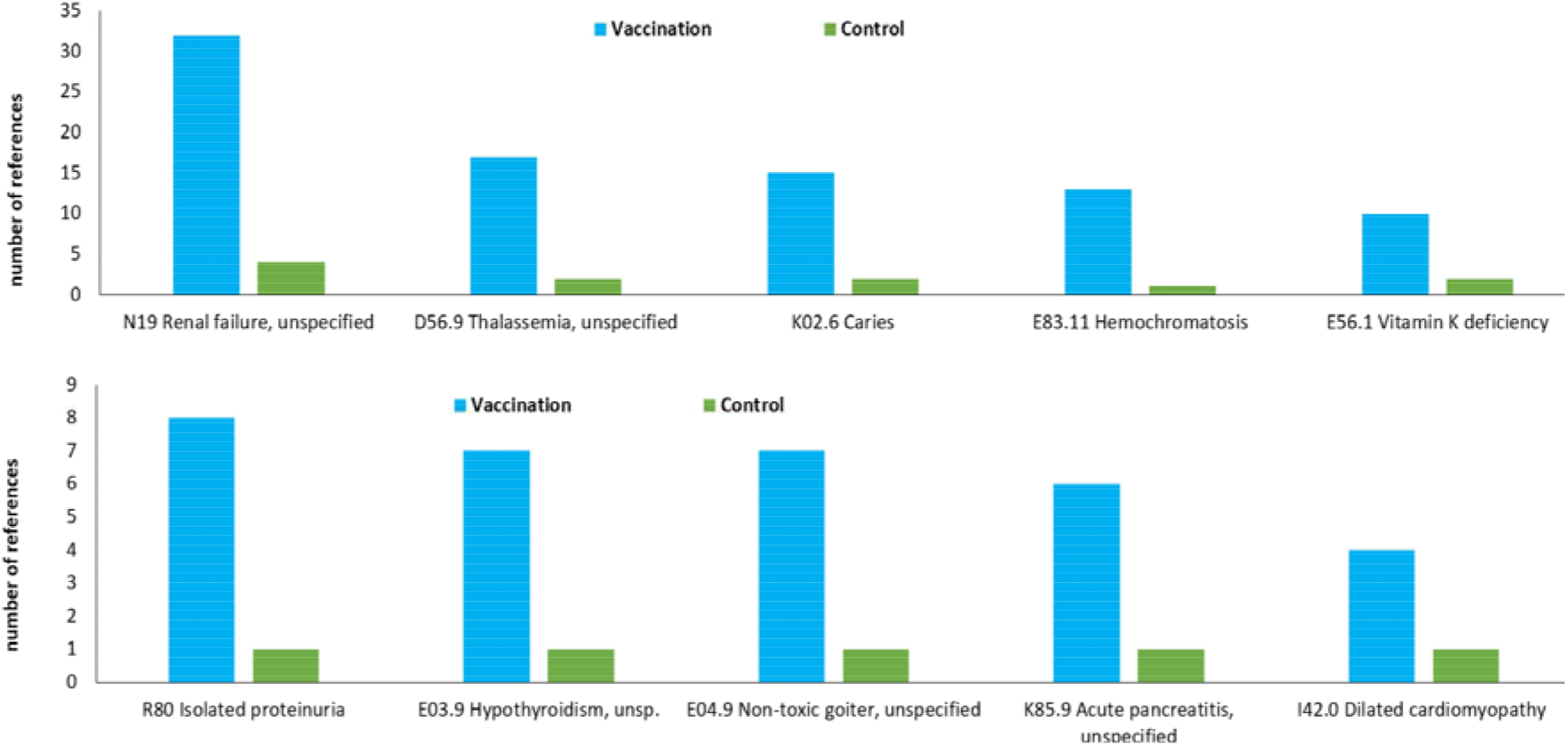
Results of mapping of the most informative terms to ICD-10 diagnoses.

Pro-inflammatory reactions provoked by vaccination, can be aggravated by micronutrient deficiencies, thus disrupting the functioning of other organ systems. In particular, mechanisms of neuronal differentiation might be disrupted (GO:0021881 Wnt-activation of neuronal differentiation, GO:0042813 Wnt-activated receptors, GO:0060828 regulation of Wnt, GO:0045813 activation of the Wnt signaling pathway, GO:1904887 Wnt signalanosome, GO:0061564 axonal development, GO:0036515 growth of serotonergic neurons), which increases the risk of neurodegenerative pathologies (Alzheimer’s disease; amyotrophic lateral sclerosis; Parkinson’s disease.), hyperalgesia, hyperactivity, autism. This topic is obviously beyond the scope of this article.

Further, we consider the relationship between the provision of various essential micronutrients and the results of vaccination against viral and bacterial pathogens.

## Discussion

### Vitamin A and vaccination

Vitamin A (retinoids) represents a hormonal factor necessary for the growth and differentiation of various cell types. The biological effects of retinoids are realized through interactions of retinoids with a number of retinoid receptors (RARA, RARB, RARG genes), including retinoid X-receptors (RXRA, RXRB, RXRG) and RAR-associated retinoid receptors (RORA, RORB, RORC). After binding to retinoid molecules, these receptors modulate the expression of several thousand target genes. RORA receptor, involved in the synthesis of cytokines, is expressed predominantly in T-cells. The activity of retinoid receptors is important for inhibition of allergic reactions, for reduction of mortality from measles, etc. [22].

The vitamin A is important for the maintenance of both cytotoxic CD8+ and T-helper CD4+ T-lymphocytes (Fig. 1). By supporting the activity of T-helpers, vitamin A also indirectly affects the activity of B-lymphocytes that produce antibodies. Thus, vitamin A deficiency can significantly disrupt the functioning of acquired immunity, which will affect the efficiency and safety of vaccination against viral and bacterial pathogens.

Vitamin A supplements have been shown to increase the titer of specific antibodies for influenza vaccination in obese mice and significantly reduced viral load after infection with the virus [23]. Oral administration of retinyl palmitate or retinoic acid to vitamin-A-deficient mice enhanced mucosal IgA antibody responses to the intranasal influenza virus vaccine [24].

Vitamin A supplementation during pregnancy enhanced maternal response to H1N1 vaccine (n = 112). The prevalence of vitamin A deficiency was very high (76%). Participants received either 10,000 IU/week vitamin A *per os* or placebo from the 2^nd^ trimester to 6 months after childbirth and were vaccinated during the 3^rd^ trimester. At 6 months postpartum, women in the vitamin A group had hemagglutination levels 38.7% higher than in the placebo group [25].

Experimental studies shown that vitamin A deficiency negatively affects results of vaccination [25-30], including usage of intranasal vaccine against RSV in newborn calves [26], intramuscular vaccination with inactivated bovine coronavirus (BCoV) [27], responses to a recombinant adenovirus vaccine in mice [28], to pentavalent rotavirus vaccine in newborn piglets [29, 30] and pneumococcal vaccine in mice [31]. Oral intake of vitamin A, on the contrary, restores the impaired immune responses thus increasing the effectiveness of the vaccination [27, 28].

Vitamin A and D supplements (25,000 IU of vitamin A and 2,500 IU of vitamin D every 15 days, totally 6 doses) improved hepatitis B vaccination outcomes in children 7-36 months of age. With vitamins, the incidence of insufficient immune response was 0% (0/37), without vitamins - 10.8% (4/37, P = 0.040) [32].

The combination of vitamins A and D (1,500 IU/day of vitamin A, 500 IU/day of vitamin D, 3 months) may improve the response to BCG vaccine in infants (n = 597). The diameter of the scars from BCG positively correlated with the diameter of the skin tightening after the Perquet test (r = 0.17, P <0.05). The frequency of positive responses of the Perquet reaction was higher in the group receiving vitamins (96.1%) than in the control group (89.7%, OR 1.07, 95% CI 1.02–1.12, P <0.05) [31]. Vitamin A supplements (15 mg plus vaccine) also increased the safety of the pentavalent diphtheria-poliomyelitis-tetanus-influenza B-hepatitis B vaccine given to infants 6, 10 and 14 weeks of age according to WHO recommendations [33].

A meta-analysis of 5 studies showed a dose-dependent effect of vitamin A supplementation on mortality after measles vaccination. Vaccination was 85% effective (95% CI 83–87). At the same time, vitamin A supplementation during the vaccination period reduced measles mortality by 62% (95% CI 19–82) [34].

Vitamin A improved the immune response (antibody levels) to an oral polio virus vaccine. Mothers in the experimental group received vitamin A (equivalent to 60 mg retinol) 3-4 weeks after delivery, and infants received 7.5 mg retinol with each vaccine dose at 6, 10 and 14 weeks of age. Vitamin A supplements increased the proportion of infants with normal titers of protective antibodies against poliovirus type 1 (RR 1.15, 95% CI 1.03–1.28) after immunization [35]. Concomitant administration of vitamin A during routine immunization also enhanced the antibody response to diphtheria vaccine in children under 6 months of age [36]. As a conclusion, vitamin A supplementation improves immune responses to vaccination against a number of viral and bacterial pathogens.

### Vitamin D and vaccination

Vitamin D, most commonly used as cholecalciferol (vitamin D3), circulates in the blood as 25-hydroxyvitamin, 25(OH)D3, from which the active, “hormonal” form 1,25(OH)2D3 is synthesized as needed to activate receptor VDR. This leads to changes in the transcription of more than 2000 human genes. The established mechanisms of the effect of vitamin D3 on immunity include the regulation of T-helper division, B-cell differentiation, modulation of cytokine profile and of the effects of interferons. For example, in response to mycobacterial infection vitamin D regulates the levels of the pro-inflammatory cytokines IL-6, TNF-alpha, and γ-interferon with participation of the Toll-like receptors TLR2, TLR4, dectin-1 and the mannose receptor (which leads, in particular, to synthesis of the antimicrobial/antiviral peptides cathelicidin and defensin) [37].

Participation of vitamin D in supporting the functioning of the cells of the acquired immunity suggests the importance of an adequate supply of vitamin D for the effectiveness and safety of vaccination against viral and bacterial pathogens. Indeed, vitamin D deficiency (serum 25(OH)D3<20 ng/ml) at initial hepatitis B vaccination was associated with lower antibody levels during the vaccination [38]. Vitamin A and D supplements help to improve the immune response to influenza vaccination in children with both vitamin deficiencies [39].

A meta-analysis of 4 vaccination studies (n = 2,367, age range 3-80 years) showed lower levels of seroprotection against influenza A/H3N2 virus when patients had vitamin D insufficiency (71.8%, the least acceptable value being 70%). In patients with normal 25(OH)D3 levels (>30 ng/ml), seroprotection was 80.1% (OR 0.63, 95% CI 0.43-0.91, p = 0.01). A similar picture was observed in the case of vaccination against the influenza B virus: against the background of vitamin D deficiency, seroprotection was only 69.6%, and with 25(OH)D3 levels above 30 ng/ml - 76.4% (OR 0.68, 95% CI 0.5-0.93, p = 0.01 [40].

It is important to emphasize that vitamin D3 has distinct antiviral effects on its own. For example, a comparative study of the efficacy and safety of the triple antiviral vaccine against measles, mumps and rubella and vitamin D3 in the treatment of warts (caused by papillomaviruses) was carried out. Patients of the first group received vaccine injections into the largest wart (n = 30), and patients of the second group received injections of vitamin D3 (n = 30) every 4 weeks, 3 times in total. Complete elimination of the largest wart was observed in 26 (87%) patients in the vaccine group and in 23 (77%) patients receiving vitamin D3. Warts distant from the injection site disappeared in 23 (77%) patients in the vaccinated group and in 20 (66%) patients in the vitamin D3 group. No significant differences were found between groups [41]. Thus, vitamin D can improve results of vaccination.

### Folates, other group B vitamins

Group B vitamins are essential for maintaining both energy metabolism and cell growth. In particular, folates, pyridoxine (vitamin B6) and cyanocobalamin (vitamin B12) are required to maintain DNA methylation, making these three vitamins essential for cell division. Folate deficiency is associated with impaired growth of blood cells (including erythrocytes and leukocytes) and impairs the immune response [42]. The influence of folates on lymphocyte growth is responsible for the effects of supplementation of folic acid on vaccination outcomes.

In particular, folic acid supplementation during pregnancy improved immunocompetence and antibody titers even 5 years after newborns were vaccinated against hepatitis B virus (n = 1461). The analysis showed an overall significant effect of folic acid intake on the increase of resistance to hepatitis B (OR 1.10, 95% CI: 1.03-1.17, p = 0.001) [43].

Supplementation with folic acid, vitamin B12 and iron during pregnancy and postpartum improves the response to influenza A virus vaccine in mothers with iron deficiency anemia (IDA, hemoglobin <110 g/L at 11-14 weeks gestation). A group of pregnant women was randomized to receive 250 μg/day B12 + 60 mg/day iron + 400 μg/day folic acid throughout pregnancy and for 3 months after delivery. The women were immunized with pandemic influenza A (H1N1) vaccine for 26-28 weeks. At the beginning of the study, 26% of women had a vitamin B12 deficiency (<150 pmol/l), 40% had vitamin B12 deficiency (150-220 pmol/l), 43% had increased methylmalonic aldehyde (MMA>271 nmol/l), 31% had a significantly increased level of homocysteine (>10 μmol/L). Vitamin supplementation increased plasma, colostrum, and breast milk B12 concentrations of mothers (p <0.05) and decreased blood MMA concentrations in mothers, newborns, and 3-month-old infants (p<0.05). Compared to placebo this vitamin-mineral supplementation significantly increased H1N1-specific IgA antibodies in plasma and colostrum of mothers and decreased the proportion of infants with elevated levels of alpha-1-acid glycoprotein and C-reactive protein [44].

### Zinc and the effects of vaccination

It is generally known that zinc ion is a cofactor of many enzymes and proteins of the human proteome. Systems biology analysis of the human proteome has shown that there are more than 1,200 Zn-binding proteins and enzymes, the activity of which is significantly reduced under conditions of zinc deficiency. More than half of these Zn-binding proteins are “zinc finger” transcription factors involved in the regulation of the expression of virtually all human genes. Disruptions in the activity of zinc-dependent signaling pathways due to zinc deficiency are associated not only with developmental anomalies and congenital endocrine pathologies but also with immunity disorders [45].

Zinc ions intensively accumulate in lymphocytes. Evidence-based studies confirm the advisability of using zinc preparations to reduce the total duration of acute respiratory diseases (influenza, adenovirus infections, etc.) and for relief of individual symptoms in a patient (runny nose, nasal congestion, sore throat, hoarseness, cough, muscle pain) [46, 47].

In an animal study, gestational zinc deficiency impaired the humoral and T-cell immune responses to hepatitis B vaccination in mice. Zinc deficiency suppressed the forcing of antibodies, decreased the ability of T-lymphocytes to divide, decreased the secretion of γ-interferon from CD4+/CD8+ T-cells [48].

Zinc deficiency in the diet reduced the humoral and T-cell immune responses to BCG vaccination in rats. Pregnant rats were divided into two groups and received a standard diet (30 mg/kg/day zinc) or a zinc-deficient diet (8 mg/kg/day zinc) for 17 weeks. Newborn rat pups were immunized with BCG vaccine (or MTB antigen ESAT-6/CFP-10) 0 and 2 weeks after birth. Adult male rats were immunized at 12 and 14 weeks of the experiment. In both pups and adults, zinc deficiency led to a decrease in the expression of Zn-transport channels ZIP2, ZIP8, and IL-6 and a decrease in rates of cell division of T-lymphocytes [49].

The effects of a zinc-deficient diet (10 mg/kg zinc) on the immune response after hepatitis B vaccination in rats were compared to effects of a zinc-sufficient diet (30 mg/kg zinc). Hepatitis B vaccine was administered intramuscularly after 8 weeks of feeding, and 4 weeks after the first injection of the “booster” dose. The mean serum zinc was 39 μg/dL (95% CI 23-75 μg/dL) on a zinc-deficient diet and 76 μg/dL (95% CI 64-115 μg/dL) on a zinc-sufficient diet (p<0.05). With zinc deficiency, an 8-fold decrease in the levels of antibodies to the hepatitis B virus (741 IU/L, 95% 0-10,000) was observed compared to zinc sufficiency (5,791 IU/L, 95% CI 558-10,000 IU/L, p<0.05) [50].

There is a correlation between the serum zinc level and the titer of antibodies produced by the tetanus vaccine in children (OR 1.84; 95% CI 1.07–3.17, p = 0.028). This association persisted regardless of age, gender, birth weight, diarrhea, breastfeeding history, serum ferritin and retinol concentrations, and malnutrition [51].

Supplementation of zinc (5 mg/day) and of a probiotic (Lactobacillus rhamnosus GG, 10^10^ CFU/day) influenced the immune response to oral rotavirus vaccine in infants aged 5 weeks. Rotavirus is the leading cause of death in children from diarrhea worldwide, and oral rotavirus vaccines are less effective than injectable ones. Two doses of the oral vaccine were given at 6 and 10 weeks of age. In children who received zinc and a probiotic, seroconversion (the percentage of individuals with a fourfold increase in antibody titer after vaccination) was 39.4% (placebo - 27.4%) [52].

### Selenium and vaccination

Selenium is required for the biosynthesis of the antioxidant glutathione and of 25 selenium-containing proteins of the human proteome [53]. Selenium supplements were shown to enhance the immune responses elicited by the avian influenza virus vaccine in chicken [54].

Supplementation with selenium-fortified yeast has been shown to benefit immunity from influenza vaccination in adults aged 50-64 years who had selenium insufficiency (<110 ng/mL in plasma). Selenium supplementation dose-dependently increased the division of T-lymphocytes (500%, SeY-50/100/200/day). When T-lymphocytes were infected with influenza viruses in culture, T-lymphocytes from patients receiving selenium were characterized by a more pronounced response of the levels of IL-8 (+169% with a Se donation of 100 μg/day) and IL-10 (+ 317%, a Se donation of 200 mg/day) [55].

Selenium supplementation affects the immune response to hepatitis B vaccine in patients with T2DM. Hepatitis B vaccine (20 mcg, days 0, 10, 21) was administered i/m after taking 200 mcg/day of selenium or placebo. Seroconversion was achieved in 23 cases (74.2%) in the selenium group and only in 15 cases (48.4%) in the placebo group (p = 0.037). Average levels of antibodies were 1233.75 ± 163.45 U/L in the selenium group and 144 ± 69.29 U/L in the control group [56].

### Iron and vaccination

Exposure to excess of lead and to iron-deficient diet affects immune response to vaccination against tetanus in rats. In particular, a significant decrease in the levels of specific IgM was noted, and the levels of T-lymphocytes of the CD8+ type were increased indicating a derangement of the T-cell-mediated mucous and humoral immune response [57].

Replenishing iron deficiency during vaccination increases the response to various types of vaccines in children (n = 303). It is important to note that hemoglobin levels during vaccination were the strongest predictor of levels of antibodies to the causative agent of diphtheria (p = 0.0071), whooping cough-IgG (p = 0.0339), pertussis filamentous hemagglutinin-IgG (p = 0.0423), and IgG to pneumococcus (p = 0.0129) in response to the use of corresponding vaccines [58].

The presence of IDA and the serum concentration of transferrin receptor during vaccination were the strongest predictors of seroconversion values after vaccination against diphtheria, pneumococcus and measles. During vaccination against measles, the effects of vitamin and iron supplements were tested (5 mg/day of elemental iron within ferrous fumarate, 4 months starting at the age of 7.5 months). The children were vaccinated against measles at 9 and 18 months. Compared to placebo 11.5 months after vaccination, children who received ferrous fumarate and other micronutrients had higher IgG antibodies to measles (p = 0.0415), higher seroconversion (p = 0.0531) and avidity (strength of cooperative antigen-antibody interactions) for IgG antibodies (p = 0.0425) [58].

### Manganese and vaccination

Manganese supplements have improved the immunocompetence of broilers following Salmonella Enteritidis vaccination. Vaccinated broilers receiving arginine and a manganese-fortified diet had higher levels of T-helper cells, T-cytotoxic, activated T-cytotoxic, and higher levels of IgM antibodies [59].

### Omega-3 polyunsaturated fatty acids and vaccination

Vaccine-induced pro-inflammatory responses are interrelated with the metabolism of pro-inflammatory prostanoids (GO:1900139 inhibition of arachidonic acid secretion, GO:0031774 leukotriene receptors, GO:0036101 leukotriene B4 catabolism, GO:0004464 leukotriene-C4 synthase etc., see Fig. 1). Omega-3 polyunsaturated acids (PUFA) such as eicosapentaenoic acid (EPA) and docosahexaenoic acid (DHA) are involved in the regulation of prostaglandin metabolism.

Omega-3 PUFA supplementation during pregnancy and lactation improves specific immunity levels induced by diphtheria and tetanus vaccinations. Pregnant women at risk of developing allergies were assigned to receive 1.6 g/day of EPA and 1.1g/day of DHA or placebo from the 25th week of gestation to 3.5 months of lactation. Omega-3 PUFA intake was associated with higher levels of the Th1-associated cytokine CXCL11 (p<0.05) and with an increase in IgG titers to diphtheria (p = 0.01) and tetanus (p = 0.05) [60].

The strength of the present research is related to a comprehensive, semi-automated analysis of all currently available texts of publications that allows to find the most relevant research on the micronutrients and vaccination. The weakness is that the dynamics of publications on the subject is quite high (in particular, due to COVID-19 research) so an adequate update will be required after some time.

In practice, the correction of micronutrient status can be done through premixes to enrich food (porridge, soup, vegetable puree, compote, juice, etc.) with vitamins and microelements. Such premixes are powdered mixtures of micronutrients containing vitamins and minerals from 50% to 100% of the physiological norms of consumption. For example, the EnzoVit Plus premix based on DSM substances provides 80% of the daily requirement for vitamins C, B1, B2, PP, B6, B9, B12, E, D, beta-carotene and minerals (iron, iodine) is currently used for the purpose of food enrichment in several countries. In accordance with the results of this analysis, a special premix or supplement, containing vitamins A, D, folates and other B vitamins, zinc, manganese, iron, selenium and omega-3 PUFAs, can be developed to improve the efficacy and safety of vaccination.

## Conclusions

Supplementation of micronutrients represents clinically efficient, safe and cost-effective intervention that allows us (1) to eliminate population-wide micronutrient deficiencies and subclinical insufficiencies, (2) to slow down the development of chronic non-communicable diseases (“diseases of civilization”), (3) to control respiratory viral infections, (4) to increase the effectiveness of various therapeutic procedures. In particular, it is advisable to use supplements/premixes of vitamins and microelements to increase the efficiency and safety of vaccination.

Currently, vaccinations against COVID-19 are being carried out in the entire world on an unprecedented scale. Both healthy people and patients with chronic conditions are vaccinated: obesity, diabetes mellitus, arterial hypertension, diseases of the liver, kidneys, etc. According to large-scale clinical and epidemiological studies, chronic comorbid pathologies at any age, especially in the elderly, are accompanied by combined deficiencies of many micronutrients [4, 6]. Hence, vaccination of such populations without micronutrient support may not only show a reduced immune response, but also provoke complications of vaccination. Preceding vaccination with micronutrient supplementation might prevent side-effects of vaccination, increase antibody titers against bacterial and viral pathogens, and reduce mortality and the severity of pathology in case of contracting the infection.

## Data Availability

The research data are available upon reasonable request

